# Impact of a team-based versus individual clinician-focused training approach on primary healthcare professionals’ intention to have serious illness conversations with patients: a theory informed process evaluation of a cluster randomized trial

**DOI:** 10.1101/2024.02.05.24302368

**Authors:** Lucas Gomes Souza, Patrick Archambault, Dalil Asmaou Bouba, Suélène Georgina Dofara, Sabrina Guay-Bélanger, Sergio Cortez Ghio, Souleymane Gadio, LeAnn Michaels, Jean-Sébastien Paquette, Shigeko (Seiko) Izumi, Annette M. Totten, France Légaré, The Meta-LARC ACP Cluster Randomized Trial team

## Abstract

**Background:** Cluster Randomized Trials (cRTs) conducted in real-world settings face complex challenges due to diverse practices and populations. Process evaluations alongside cRTs can help explain their results by exploring possible causal mechanisms as the trial proceeds.

**Objective:** To conduct a process evaluation alongside a cRT that compared the impact of team-based vs. individual clinician-focused SICP training on primary healthcare professionals’ (PHCPs) intention to have serious illness conversations with patients.

**Methods:** The cRT involved 45 primary care practices randomized into a team-based (intervention) or individual clinician-focused training program (comparator) and measured primary outcomes at the patient level: days at home and goal of care. Our theory-informed mixed-methods process evaluation alongside the cRT measured intention to have serious illness conversations with patients among the trained PHCPs using the CPD-Reaction tool. Barriers and facilitators to implementing serious illness conversations were identified through open-ended questions and analyzed using the Theoretical Domains Framework. We used the COM-B framework to perform triangulation of data. We reported results using the CONSORT and GRAMMS reporting guidelines.

**Results:** Of 535 PHCPs from 45 practices, 373 (69.7%) fully completed CPD-Reaction (30.8% between 25-34 years old; 78.0% women; 54.2% had a doctoral degree; 50.1% were primary care physicians). Mean intention scores for the team-based (n=223) and individual clinician-focused arms (n=150) were 5.97 (Standard Error: 0.11) and 6.42 (Standard Error: 0.13), respectively. Mean difference between arms was 0.0 (95% CI −0.30;0.29; p=0.99) after adjusting for age, education and profession. The team-based arm reported barriers with communication, workflow, and more discomfort in having serious illness conversations with patients.

**Conclusions:** Team-based training did not outperform individual clinician-focused in influencing PHCPs’ intention to have serious illness conversations. Future team-based interventions could foster behaviour adoption by focusing on interprofessional communication, better organized workflows, and better support and training for non-clinician team members.

**Registration:** ClinicalTrials.gov (ID: NCT03577002).

## INTRODUCTION

Cluster Randomized Trials (cRTs) using a pragmatic approach aim to evaluate the effectiveness of interventions in real-world clinical settings, providing advantages such as enhanced generalizability and relevance to routine care (1). However, these cRTs present unique challenges, particularly in terms of implementation. Implementing and maintaining interventions in cRTs within the complex environment of routine clinical care proves difficult due to variations in clinical practices, healthcare systems, and participant populations (2). Therefore, process evaluations alongside cRTs are an essential tool and are increasingly used to shed light on the ‘black box’ of complex interventions and to provide information to interpret outcome results and aid moving them into practice (3, 4).

In 2017, the Meta-Network Learning and Research Center Advanced Care Planning (Meta-LARC ACP) cRT team designed a comparative effectiveness cRT to assess two training approaches for the serious illness care program (SICP) developed by Ariadne Labs (5, 6). The cRT compared a team-based training approach to conducting SICP (intervention) with the traditional clinician-based training approach (comparator) (7). The rationale for this cRT stemmed from the initial assumption in the design of the SICP which presumed that only individual clinicians, such as physicians, nurse practitioners, or physician assistants, would be responsible alone for having serious illness conversations with patients. However, insights drawn from studies on team approaches to chronic and complex care suggest that adopting a team-based approach could facilitate the implementation of serious illness conversations within clinical practice (8, 9).

In recognition of the pivotal role played by primary care teams in patient care, the cRT explored a training approach that integrated task-sharing strategies. These strategies encompassed patient identification, conversation preparation, discussion initiation, and follow-up (8, 10). This training approach aligned with systematic reviews which identified time constraints as a significant barrier to implementing these discussions in primary care settings. The involvement of multiple primary healthcare professionals (PHCPs) - primary care clinicians and other primary care team members - could potentially alleviate this burden by reducing the time commitment required from each individual, thereby facilitating effective implementation (11, 12). Despite the promising potential of a team-based SICP training approach, evidence supporting its efficacy and implementation in primary care was limited at the time of the cRT’s conception (13). This underscored the need for its evaluation in a real-world primary care setting.

To gain a more in-depth understanding of the intervention implementation process, which was the team-based SICP training approach, we conducted a process evaluation alongside the parent cRT by comparing the intervention’s impact on PHCPs’ instead of patients, and specifically, on their intention to have serious illness conversations with patients. Process evaluations alongside cRTs aim to provide insights into how an intervention was delivered, how participants received it, and whether it was implemented as intended. Our process evaluation aimed to help us understand PHCP uptake of the training and their intention to have serious illness conversations with patients. This process evaluation was also intended to identify factors influencing PHCPs’ intention to have serious illness conversation with patients, such as the modifiable psychosocial factors influencing their intention and the perceived barriers and facilitators to adopting the target behaviour. In order to achieve these aims, we incorporated established theories of behaviour change into our process evaluation (14). These theoretical frameworks played a key role in identifying and addressing specific implementation challenges, enhancing our understanding of the intervention, and suggesting modifications to optimize its impact in future interventions (15). Our process evaluation is particularly informed by a set of social cognitive theories. First, the integrated model of behaviour change in healthcare professionals, informed by a systematic review of 76 studies, identifies intention as a mediator of behaviour and influenced by six factors: capability, consequence, moral norm, social influence, role/identity, and healthcare professional characteristics (16). The Theoretical Domains Framework (TDF) expands on this by identifying additional factors influencing healthcare professionals’ behaviour related to implementing recommendations (17). Finally, the COM-B model maps these factors onto specific behavior change techniques to guide future intervention development (18).

We hypothesized, based on our previous theory-informed studies on interprofessional interventions (19, 20), that team-based training would outperform individual clinician-focused training in influencing PHCPs intention to have serious illness conversations with their patients. These studies demonstrated that our interprofessional model effectively promoted positive intentions. Furthermore, the research elucidated the modifiable psychosocial factors that positively correlate with intention within an interprofessional context (19, 20). Therefore, we performed a theory-based process evaluation alongside our parent cRT to assess the effect of the two SICP training approaches on PHCPs’ intention to have serious illness conversations with patients and to identify potential facilitators and barriers to the adoption of this behaviour.

## MATERIALS AND METHODS

### Study design and setting

To assess the implementation of a team-based SICP training approach compared to an individual clinician-focused SICP training approach, we conducted a process evaluation alongside a parent cRT that used a comparative effectiveness design (21, 22). This process evaluation employed a mixed-methods concurrent embedded design (23), utilizing secondary post-intervention qualitative and quantitative data from the parent cRT. Process evaluations often employ mixed-methods designs due to their ability to provide a more comprehensive and multifaceted understanding of intervention implementation, encompassing both quantitative and qualitative data(15, 24). We used Godin et al. integrative model of behavior prediction in healthcare professionals for our quantitative analysis (16, 25, 26), the Theoretical Domains Framework (TDF) for our qualitative analysis (17, 27), and the Capability, Opportunity, Motivation and Behaviour (COM-B) model to triangulate findings (28). We reported this paper following the extension of Consolidated Standards of Reporting Trials (CONSORT) for cRTs (S1 Checklist) and followed the Good Reporting of a Mixed Methods Study (GRAMMS) checklist (S2 Checklist) (29, 30). The parent cRT and object of our process evaluation is registered at ClinicalTrials.gov (ID: NCT03577002). The protocol is published elsewhere (7) and the results for the primary outcomes at the patient level (days spent at home and goal concordant care) are under review.

The parent Meta-LARC ACP cRT was conducted in community-based primary care practices in five US states (Colorado, Iowa, North Carolina, Oregon, Wisconsin) and two Canadian provinces (Quebec and Ontario) recruited through Meta-LARC, a consortium of practice-based research networks (PBRNs) (7). A cRT design was selected as the knowledge learned in the SICP trainings was to be implemented at a practice level.

For our process evaluation study, we used the Medical Research Council framework (24) to delve into the context and mechanisms of impact of our intervention, which are key functions of a process evaluation according to this framework. We did that by primarily assessing what influenced PHCPs’ intention to adopt the target behaviour, which was defined as *having serious illness conversations with patients*. A previous study focused on the development of the intervention (31). A subsequent study will explore the contextual factors that influenced the sustainability of the intervention one year and two years after the training, particularly in the context of the COVID-19 pandemic. This approach will allow us to address all the different aspects of process evaluations as suggested by Moore et al. (24)

### Ethical approval

The parent META-LARC ACP cRT was approved by the Trial Innovation Network Single IRB at Vanderbilt University Medical Center (IRB#181084) for the U.S. sites; by the Research Ethics Board of the Centre Intégré Universitaire de Santé et de services sociaux (CIUSSS) de la Capitale-Nationale in Quebec City, Canada (ethics number #MP-13-2019-1526), for the sites in Quebec and by the Health Sciences Research Ethics Board of the University of Toronto (protocol number 36631), for the sites in Ontario. The process evaluation’s data and outcomes were included in the approval. All subjects willingly agreed to take part in the study, and their consent (verbal or written) was obtained and registered by PBRNs in accordance with the regulations of the Institutional Review Board or Research Ethics Board in effect.

### Participants and eligibility

To participate in the parent cRT a practice had to a) be willing and able to be randomized to either the team-based or individual-clinician-based SICP approach, b) have sufficient staff to participate in the team-based arm, and c) not be engaged in another standardized ACP program. Detailed criteria for practice eligibility are published elsewhere (7). In this process evaluation, we analyzed a secondary outcome, the impact of the intervention on PHCPs’ behavioural intention, and therefore participants for this process evaluation were the PHCPs (primary care clinicians and other primary care team members) working in the primary care practices enrolled in the parent cRT. PHCPs from participating eligible practices were invited to participate in the training and to answer the after-training questionnaires, but not required to participate.

### Randomization

The units of randomization were the primary care practices stratified by PBRN. To assure allocation concealment, involvement in randomization was limited to statisticians not involved in other aspects of the project. Staff at the PBRNs, practices, and the Meta-LARC coordinating center were not involved in the randomization. Statisticians completing the analysis were blinded to allocation until the parent cRT primary outcomes analysis was completed. Investigators, PBRN leadership, practices, and research staff were not blinded to the assignment. Practices and participating PHCPs could not be blinded to which approach they were assigned, as they needed to actively to be trained and implement the intervention or comparator. More details regarding randomization are published elsewhere (7).

### Intervention and comparator arms

The SICP developed by Ariadne Labs was adapted to be used by interprofessional teams of PHCPs (5). Training lasted three hours per arm: a 1.5-hour online module (Part A), and a 1.5-hour in-person role-play session (Part B) (Fig 1). Training materials are available at https://primarycareacp.org, including the Serious Illness Conversations Guide (SICG), a tool designed by the original developers of the SICP to facilitate communication with patients with serious illnesses(5).

**Fig 1.**
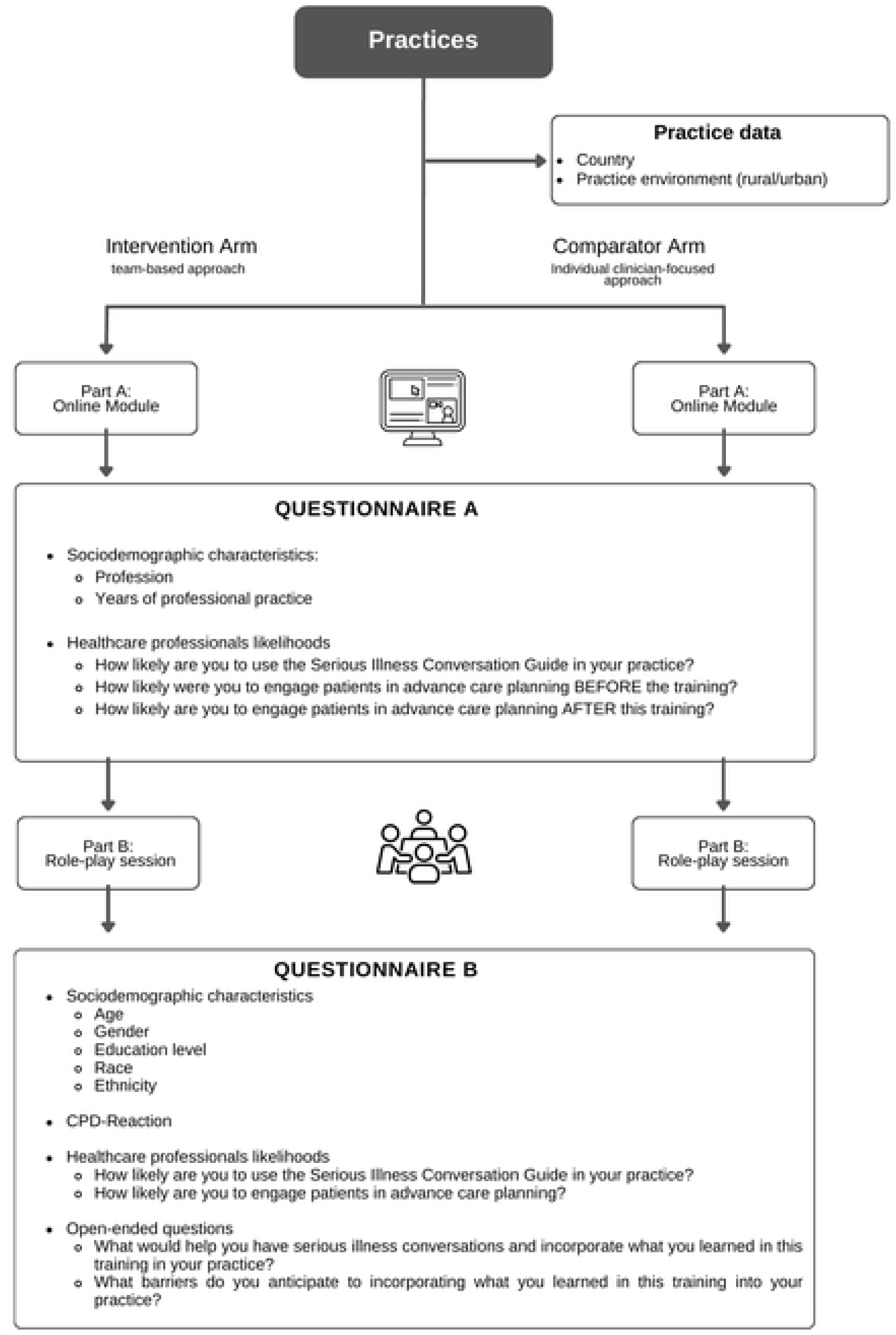
Study design and data collection

Training took place between October 2018 and November 2019. A train-the-trainer model was used whereby 1 to 3 trainers were identified by each PBRN and trained simultaneously by the core trainers. Trainers from each PBRN then trained PHCPs in their local practices. The intervention proceeded as described in the study protocol (7).

#### Intervention arm

The intervention arm received an SICP training session adapted to a team-based approach, whereby primary care clinicians (i.e., primary care physicians, physician assistants, nurse practitioners, and medical residents) and all other primary care team members, such as medical assistants, nurses, dieticians and social workers were invited to complete the training. The adapted team-based approach was based on an interprofessional shared decision-making model and workshop (32, 33) that has been used in a variety of contexts (34) and showed a positive impact on behavioural intention (35). In this training, the team received guidance on establishing a common understanding of the processes and goals of serious illness conversations, dividing and sharing tasks, recognizing each member’s contributions, promoting continuous communication in the team, and recognizing the organizational or functional constraints of each profession (31).

#### Comparator arm

The comparator arm received the individual clinician-focused approach as designed by Ariadne Labs (5). It focuses only on primary care clinicians (i.e., primary care physicians, physician assistants, nurse practitioners, and medical residents). Other primary care team members, such as medical assistants, nurses, dieticians and social workers did not participate in the training. The individual clinician-focused approach assumes that primary care clinicians alone are responsible for selecting patients, conducting, and documenting serious illness conversations.

### Outcomes and measurements

The outcome of interest for this process evaluation was to measure the impact of our training intervention on PHCPs’ intention to have serious illness conversations with patients. This focus on intention was based on its established role as a proxy for behaviour, with a demonstrated predictive capacity for behavior change up to six months following training interventions (16, 26, 36, 37). Additionally, measuring intention allowed for a targeted assessment of the intervention’s influence on PHCPs, the primary implementers of the intervention in the parent cRT. Moreover, it facilitated the identification of modifiable psychosocial factors within the real-world practice context that could be targeted to promote the adoption of the desired behaviour (16, 26, 36). Similarly, tools that measure the intention as a proxy to predict clinical behaviour are valuable for evaluating the efficacy of professional development training interventions (36). They facilitate the immediate evaluation of an intervention’s impact since evaluating behaviour in a real clinical context can be complex and costly (38). This information provides crucial data for our process evaluation, enabling the identification of intervention strengths and weaknesses related to behaviour change. By understanding these mechanisms of action, we can contribute to a cumulative implementation science knowledge base and inform the interpretation and adaptation of future SICP interventions (15).

We used the CPD-Reaction tool to measure PHCPs’ behavioural intention to have serious illness conversations with patients. CPD-Reaction is a self-administered, theory-informed, validated tool that assesses five constructs (intention, beliefs about capabilities, beliefs about consequences, moral norm, and social influences) using a 12-item self-administered questionnaire (36, 39). Most items are measured using a Likert scale of 1–7, where 1 represents “strongly disagree” and 7 represents “strongly agree.” One item, the social influences construct, ranges from 1 to 5.(25) Even though variables measured using a Likert scale are categorical, intention can be analyzed as a continuous variable with no harm to the analysis (40).

However, as proposed by Lou, Atkins and West (17), behaviours are a part of an intricate system, they do not occur in isolation. Consequently by incorporating and systematically expanding the range of relevant behaviours measured, while maintaining appropriate selectivity, we were able to increase both the depth and breadth of our analysis (18). Therefore, to gain a more comprehensive understanding of the intervention’s potential impact, we decided to measure not only PHCPs intention to have serious illness conversations with patients (the primary outcome), but also their likelihood to engage in ACP and use the SICG as additional outcomes for this process evaluation. We chose likelihood questions for these additional outcomes because they are simple, straightforward, and are a widely used tool in the literature to capture intentions and consequently the concurrent behaviours we wanted to measure (41). The choice of a simpler one-item type of question had the aim to optimize participant engagement and data completeness and to minimize survey fatigue among PHCPs. Thus, after Part A of the training session, we asked PHCPs about what their likelihood to engage in ACP was, both before and after Part A, using a retrospective pretest-posttest evaluation design (42). Then, after Part B, we also asked how likely they were to use the SICG in their practices. Face and content validity of the likelihood measures were established based on studies on similar training programs with PHCPs (43, 44). These questions also used a Likert scale of 1-10 where 1 represented “extremely unlikely”, 5 represented “moderate” and 10 “extremely likely”. In S1 Fig, we present a diagram illustrating the interactions among the behaviours we measured, offering a structured perspective on their interplay.

Finally, using two open-ended questions we evaluated the facilitators and barriers to PHCPs incorporating the knowledge acquired in their practices and to have serious illness conversations with patients. We mapped these facilitators and barriers onto the TDF. The TDF was developed through a consensus of experts who consolidated 33 psychosocial theories of behaviour change to generate 14 domains (45). Fig 1. details all the process evaluation measurements and outcomes sought throughout the different steps of the intervention.

### Data collection

Data collection took place between October 2018 and November 2019. Before randomization, sociodemographic data were collected as well as additional data directly from practices (e.g., country and practice environment) (Fig. 1). This analysis uses the data collected through questionnaires administered immediately after training was completed in each practice, as we were interested in PHCPs’ immediate evaluation after the training. This helped us understand if the team-based SICP training approach was effective in impacting PHCPs’ intention compared to the individual clinician-focused SICP training and consequently their behavior, thus helping us understand the mechanisms of impact of our intervention (24). To optimize the assessment and maximize the time of participating PHCPs, we employed self-administered questionnaires in the study. We also opted for open-ended questions instead of conducting semi-structured interviews or focus groups to explore barriers and facilitators. These decisions streamlined data collection, enhancing both efficiency and participant convenience.

### Sample size

The sample size calculation for the parent study is available in the published protocol (7). A sample size estimate for PHCPs involved in our process evaluation study was not calculated as their responses were not the primary outcome of the parent cRT A total of 535 PHCPs attended the trainings, 326 in the team-based and 209 in the individual clinician-focused arms. We analyzed data from 373 PHCPs (223 team-based and 150 individual clinician-focused) who attended the training and completed the intention construct in the CPD-Reaction tool. For these 373 PHCPs, we obtained a post-hoc power of 0.75 based on an unadjusted mixed linear regression model.

### Analysis

#### Quantitative analysis

We used descriptive statistics to report our variables including practice and participant characteristics. Categorical variables were described with absolute (n) and relative (%) frequencies; and continuous variables with their central tendency measures (mean) and dispersion (standard deviation).

Then, we used a mixed linear model to compare mean scores between study arms for PHCPs’ intention to have serious illness conversations with patients. To account for clustering, we used the practice identifier as a random factor when fitting the models.

Next, we fit a bivariate mixed linear model for each sociodemographic variable of interest to examine its effect on PHCPs’ intention to have serious illness conversations with patients. We subsequently fit a multivariable mixed linear model including all variables for which we detected a potential effect (p<0.20) on PHCPs’ intention in the bivariate analyses. We then conducted manual backward stepwise selection based on variable significance for a final adjusted model. The arm variable was kept in the model regardless of significance as our objective was to compare the impact of training approaches.

Finally, we fit mixed linear models to evaluate how likely participants considered they were to engage patients in ACP (i.e. before, reported retrospectively, and then after training), and how likely they were to use the SICG in their practice after both parts of the training sessions. We compared the mean difference between the likelihood to engage in ACP measured before and after Part A of the training between each arm, and also regarding their likelihood to use the SIC Guide, also measured between each arm. We used the practice identifier as a random factor in all models. All analyses were performed with SAS (Statistical Analysis Software) 9.4.

#### Qualitative analysis

To further explore intention, using the TDF as a guide, we performed a descriptive analysis of the answers to the two open-ended questions (17, 46). Two researchers (SGD, LGS) used an inductive approach to develop the codes that were used in the thematic analysis (47). Then, two researchers (LGS, DAB) conducted a thematic analysis of the answers, and any disagreements between the researchers were resolved by consensus. If the disagreement remained a third senior researcher was consulted (FL). Data were then deductively classified into TDF domains. We calculated the frequency of each barrier and facilitator by recording the number of times it was mentioned in the answers.

#### Triangulating qualitative and quantitative data

Our process evaluation equally intended to propose practical theory-driven recommendations to improve the intervention. To inform these recommendations, we triangulated qualitative and quantitative data to grasp a broader understanding of the psychosocial factors influencing our targeted behaviour (48). We conducted a comparison between the five psychosocial determinants assessed in the CPD-Reaction questionnaire and the domains outlined in the TDF (LGS and SGD). Our analysis involved identifying points of convergence and divergence between quantitative and qualitative data, exploring instances where both types of data provided additional insights into the same constructs. Subsequently, recommendations were formulated utilizing the COM-B model of behaviour, which posits three criteria—capacity, opportunity, motivation—for the occurrence of a behavior (18, 49). These criteria and their subcategories were linked to the TDF domains, along with their associated barriers or facilitators (S2 Fig)(50). The COM-B model further proposes nine intervention functions, aligned with TDF domains, that are capable of promoting behaviour change, namely education, persuasion, incentivization, coercion, training, restriction, environmental restructuring, modeling, and enablement (18, 28, 51, 52). By discerning which intervention functions corresponded to our results, we identified and selected behaviour change techniques associated with the relevant functions to derive our recommendations (18). The proposed recommendations were reviewed by all authors.

## RESULTS

### Practice and PHCP characteristics and flow diagram

As the parent trial is a cRT in table 1 we provide a comprehensive description of the characteristics of practices randomized Indicating that the randomization of clinics was adequate.

**Table 1:** Participating primary care practices characteristics.

|  | Total | Team-based<br>arm | Individual clinician-focused arm |
| --- | --- | --- | --- |
| **Number of practices** | 45 | 23 | 22 |
| **Country, n (%)** |  |  |  |
| United States | 33 (73.3) | 17 (73.9) | 16 (72.7) |
| Canada | 12 (26.7) | 6 (26.1) | 6 (27.3) |
| **Size (# of Primary healthcare professionals), n (%)** |  |  |  |
| Small (2-5) | 8 (17.8) | 6 (26.1) | 2 (9.0) |
| Medium (6-12) | 19 (42.2) | 9 (39.1) | 10 (45.5) |
| Large (13-85) | 18 (40.0) | 8 (34.8) | 10 (45.5) |
| **Geographic Setting, n (%)** |  |  |  |
| Rural | 20 (44.4) | 12 (52.2) | 8 (36.4) |
| Suburban | 8 (17.8) | 3 (13.0) | 5 (22.7) |
| Urban | 17 (37.8) | 8 (34.8) | 9 (40.9) |
| **Ownership, n (%)** |  |  |  |
| Hospital/health system | 31 (68.9%) | 13 (56.6) | 18 (82.8) |
| Physician or physician group | 11 (24.4) | 7 (30.4) | 4 (18.2) |
| Federally qualified health centre | 3 (6.7%) | 3 (13.0) | - |
| **Specialty, n (%)** |  |  |  |
| Family Medicine | 34 (75.5) | 15 (68.2) | 19 (82.6) |
| Internal Medicine | 8 (17.8) | 5 (22.7) | 3 (13.) |
| Both family and internal medicine | 3 (6.7) | 2 (9.1) | 1 (4.4) |
| Size of primary healthcare professionals,<br>median (min to max) | 10 (3 to 46) | 8 (4 to 46) | 12 (3 to 40) |

Fig. 2 illustrates the flow of practices and PHCPs in this process evaluation study alongside the Meta-LARC ACP cRT. Thirty-eight practices participated (19 in each arm), a participation rate of 84.4% (38/45). A total of 373 (69.7%) PHCPs fully completed the intention construct in CPD-Reaction. The sociodemographic characteristics of the participating PHCPs are detailed in Table 2.

**Fig. 2:**
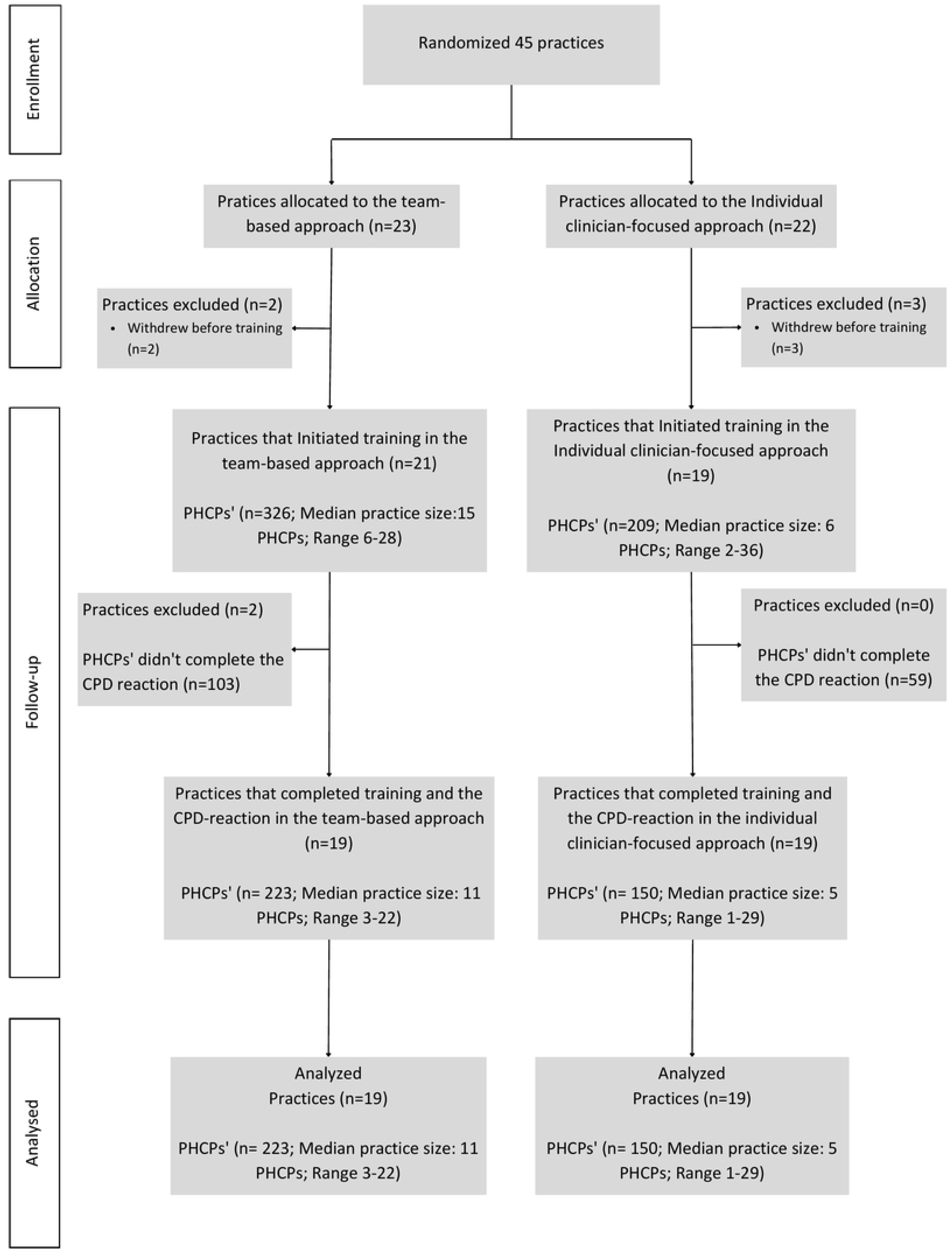
Flowchart of Primary Healthcare professionals (PHCPs)

**Table 2:** Characteristics of participating PHCPs by arm.

|  | **Total** | **Team-based arm** | **Individual clinician-**<br>**focused arm** |
| --- | --- | --- | --- |
| **Number of professionals** | 373 | 223 | 150 |
| **Country, N (%)** |  |  |  |
| USA | 216 (57.9) | 143 (64.1) | 73 (48.7) |
| Canada | 157(42.1) | 80 (35.9) | 77 (51.3) |
| **Age, N (%)** |  |  |  |
| Missing data | 12 (3.2) | 10 (4.5) | 2 (1.3) |
| 18-24 | 11 (2.9) | 8 (3.6) | 3 (2.0) |
| 25-34 | 115 (30.8) | 64 (28.7) | 51 (34.0) |
| 35-44 | 113 (30.3) | 63 (28.2) | 50 (33.3) |
| 45-54 | 64 (17.2) | 38 (17.0) | 26 (17.3) |
| 55-64 | 42 (11.3) | 30 (13.5) | 12 (8.0) |
| > 65 | 12 (3.2) | 10 (4.5) | 2 (1.3) |
| Prefer not to answer | 4 (1.0) | - | 4 (2.7) |
| **Sex, N (%)** |  |  |  |
| Missing data | 3 (0.8) | 3 (1.3) | - |
| Male | 79 (21.2) | 35 (15.7) | 44 (29.3) |
| Female | 291 (78.0) | 185 (83.0) | 106 (70.7) |
| **Ethnicity, N (%)** |  |  |  |
| Missing data | 7 (1.9) | 6 (2.7) | 1 (0.7) |
| Yes, Hispanic or Latino | 13 (3.5) | 9 (4.0) | 4 (2.7) |
| No, not Hispanic or Latino | 347 (93.0) | 205 (92.0) | 142 (94.7) |
| Prefer not to answer | 6 (1.6) | 3 (1.3) | 3 (2.0) |
| **Race, N (%)** |  |  |  |
| Missing data | 6 (1.6) | 5 (2.2) | 1 (0.7) |
| White | 320 (85.8) | 192 (86.1) | 128 (85.3) |
| Asian | 20 (5.4) | 13 (5.8) | 7 (4.7) |
| American Indian or Alaska native | 2 (0.5) | 1 (0.4) | 1 (0.7) |
| Black or African American | 14 (3.7) | 9 (4.0) | 1 (0.7) |
| Hawaiian Native or Pacific Islander | 1 (0.3) | 1 (0.4) | - |
| Others | 2 (0.5) | 1 (0.4) | 1 (0.7) |
| Prefer not to answer | 8 (2.1) | 1 (0.4) | 7 (4.7) |
| **Education level, N (%)** |  |  |  |
| Missing data | 4 (1.0) | 3 (1.4) | 1 (0.7) |
| Doctoral degree | 202 (54.2) | 91 (40.8) | 111 (74.0) |
| Master's degree | 65 (17.4) | 34 (15.2) | 31 (20.7) |
| Bachelor's degree | 40 (10.7) | 40 (17.9) | - |
| High school | 9 (2.4) | 9 (4.0) | - |
| Others | 53 (14.2) | 46 (20.6) | 7 (4.7) |
| **Profession, N (%)** |  |  |  |
| Missing data | - | - | - |
| Primary care physician | 187 (50.1) | 86 (38.6) | 101 (67.3) |
| Nurse practitioner | 35 (9.4) | 16 (7.2) | 19 (12.7) |
| Nurse | 54 (14.5) | 54 (24.2) | - |
| Physician assistant | 16 (4.3) | 7 (3.1) | 9 (6.0) |
| Social worker | 6 (1.6) | 6 (2.7) | - |
| Medical assistant | 33 (8.9) | 33 (14.9) | - |
| Resident | 29 (7.8) | 8 (3.6) | 21 (14.0) |
| Others (Psychologist, Dietician, etc.) | 13 (3.5) | 13 (5.8) | - |

### Quantitative results

#### Behavioural intention

Table 3 shows the scores for behavioural intention and its psychosocial determinants. On a scale of 1–7, the mean intention score was 5.97 (Standard error: 0.11) for the team-based arm (intervention) and 6.42 (Standard error: 0.13) for the individual clinician-focused arm (comparator). The difference in mean intention scores and 95% confidence interval (95% CI) between arms was −0.45 (−0.79 to −0.11), with a P-value of 0.01 **(Table 4)**. There was no statistically significant difference in the other CPD-Reaction constructs between arms.

**Table 3:** Comparison of intention scores and and its psychosocial determinants measured with the CPD-Reaction questionnaire after the training.

| Intention and psychosocial determinants– Score Range (1 to 7)<sup>a</sup> | Team-based arm |  | Individual clinician-focused arm |  | Estimate difference (95% CI)<sup>b</sup> | P-Value |
| --- | --- | --- | --- | --- | --- | --- |
|  | N | Mean (SE) | N | Mean (SE) |  |  |
| Beliefs about capabilities | 223 | 5.40 (0.08) | 150 | 5.60 (0.09) | -0.20 (-0.45; 0.05) | 0.11 |
| Social Influences | 217 | 4.69 (0.13) | 147 | 4.83 (0.15) | -0.13 (-0.53; 0.26) | 0.50 |
| Beliefs about consequences | 223 | 6.36 (0.07) | 150 | 6.56 (0.09) | -0.21 (-0.43; 0.02) | 0.08 |
| Moral Norm | 223 | 6.71 (0.04) | 150 | 6.81 (0.05) | -0.10 (-0.23; 0.02) | 0.10 |
| Intention | 223 | 5.97 (0.11) | 150 | 6.42 (0.13) | -0.45 (-0.79; -0.11) | **0.01** |

Analyzed using a Linear mixed model
P-value < 0.05 – statistically significant (T-test);
95% CI, confidence interval at 95%; SE, standard error of the mean
<sup>a</sup>The social influences construct is the only construct that has one item with a Likert-scale that ranges from 1 to 5.
<sup>b</sup> Least squares mean

**Table 4:** Comparison of the intention to have serious illness conversations with patients between the team-based arm and the individual clinician-focused arm.

|  | Unadjusted Model |  | Adjusted model<sup>b</sup> |  |
| --- | --- | --- | --- | --- |
|  | Mean difference (95% CI)<sup>c</sup> | P-Value<sup>a</sup> | Mean difference (95% CI)<sup>c</sup> | P-Value<sup>a</sup> |
| Individual clinician-focused arm | Reference |  | Reference |  |
| Team-based arm | -0.45 (-0.79; -0.11) | **0.01** | 0.00 (-0.30;0.29) | 0.99 |

Analyzed using a Linear mixed model
<sup>a</sup> P-value < 0.05 – statistically significant (T-test)
<sup>b</sup> Adjusted for age, profession and education level,
<sup>c</sup>95% CI, confidence interval at 95%.

However, after adjusting for education and profession, the difference in intention between the two groups was no longer statistically significant: 0.00 (95% CI: −0.30 to 0.29), with a P-value of 0.99 **(Table 4).**

#### Likelihood to engage in ACP and to use the SICG

The mean difference in scores between the two arms for “*How likely were you to engage patients in ACP before training?*” was −1.16; P<0.01, and for “*How likely are you to engage patients in ACP after training?*” was −0.68 (P=0.01) (S1 Table). Mean difference between pre- and post-training likelihood to engage in ACP was 2.95 (P<0.001) in the team-based arm and 2.60 (P<0.001) in the individual clinician-focused arm **(Table 5),** the difference between arms was not statistically significant (P=0.31). Likewise, mean difference between PHCPs’ self-reported likelihood to use the SICG in their practice after Part A and after Part B was 0.46 (P=0.01) in the team-based arm and 0.50 (P=0.02) in the individual clinician-focused arm, the difference between arms was also not statistically significant (P=0.88) **(Table 5).**

**Table 5:**
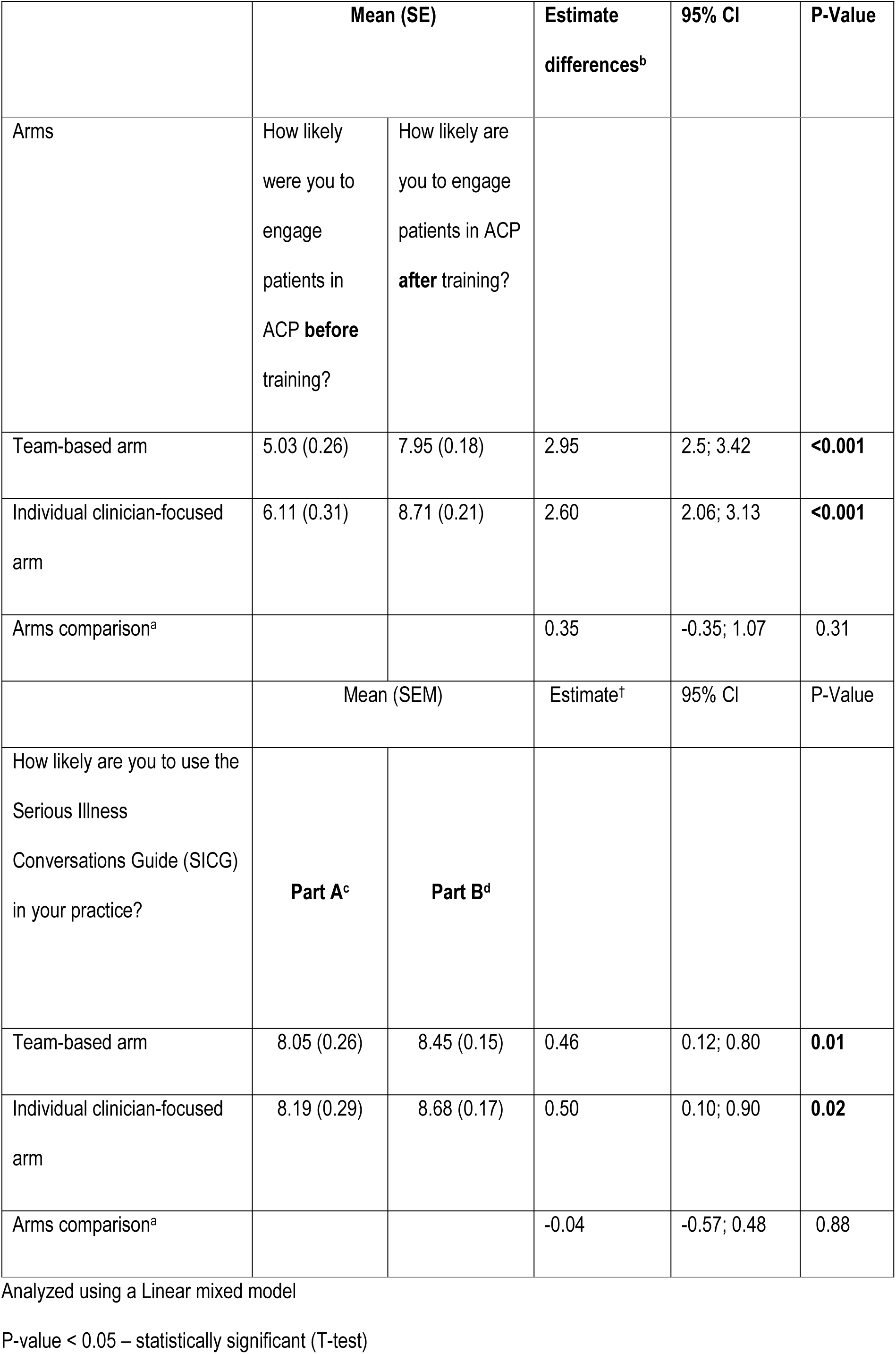
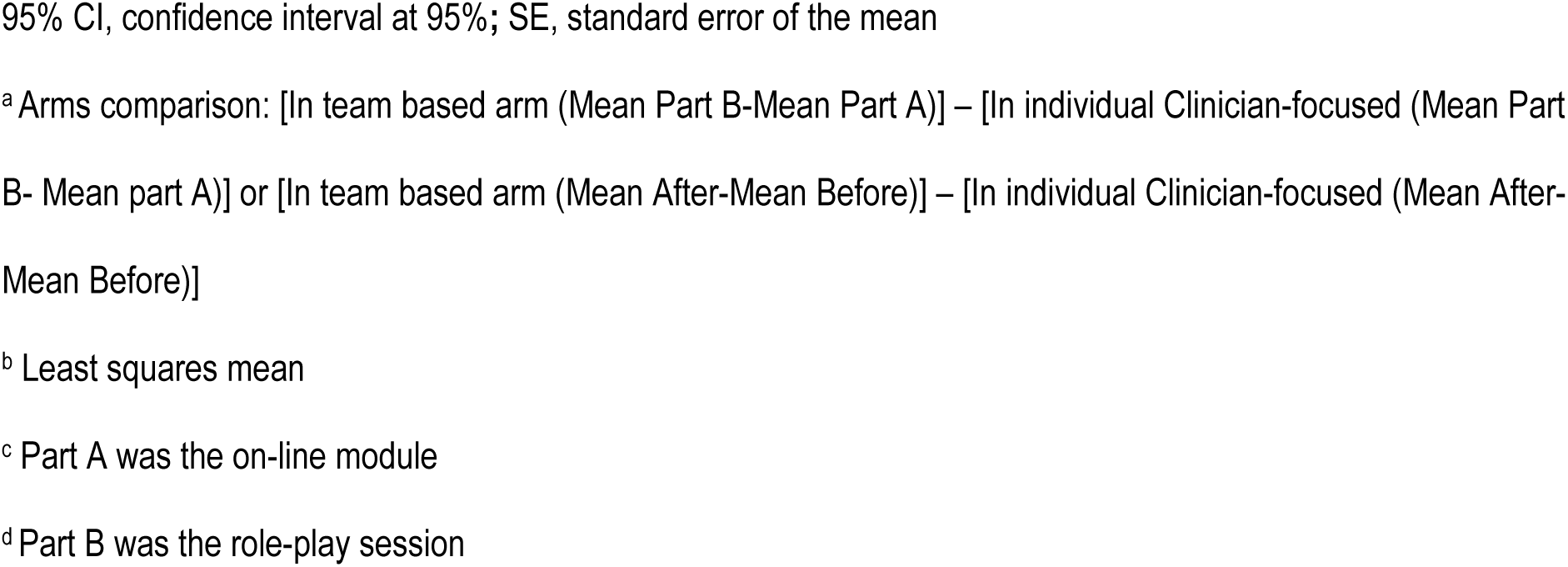
Differences in the likelihood questions about engaging patients in ACP before and after training and using the SICG.

|  | Mean (SE) |  | Estimate differences<sup>b</sup> | 95% CI | P-Value |
| --- | --- | --- | --- | --- | --- |
| Arms | How likely were you to engage patients in ACP **before** training? | How likely are you to engage patients in ACP **after** training? |  |  |  |
| Team-based arm | 5.03 (0.26) | 7.95 (0.18) | 2.95 | 2.5; 3.42 | **<0.001** |
| Individual clinician-focused arm | 6.11 (0.31) | 8.71 (0.21) | 2.60 | 2.06; 3.13 | **<0.001** |
| Arms comparison<sup>a</sup> |  |  | 0.35 | -0.35; 1.07 | 0.31 |
|  | Mean (SEM) |  | Estimate<sup>†</sup> | 95% CI | P-Value |
| How likely are you to use the Serious Illness Conversations Guide (SICG) in your practice? | **Part A<sup>c</sup>** | **Part B<sup>d</sup>** |  |  |  |
| Team-based arm | 8.05 (0.26) | 8.45 (0.15) | 0.46 | 0.12; 0.80 | **0.01** |
| Individual clinician-focused arm | 8.19 (0.29) | 8.68 (0.17) | 0.50 | 0.10; 0.90 | **0.02** |
| Arms comparison<sup>a</sup> |  |  | -0.04 | -0.57; 0.48 | 0.88 |

Analyzed using a Linear mixed model
P-value < 0.05 – statistically significant (T-test)
95% CI, confidence interval at 95%; SE, standard error of the mean
<sup>a</sup> Arms comparison: [In team based arm (Mean Part B-Mean Part A)] – [In individual Clinician-focused (Mean Part B- Mean part A)] or [In team based arm (Mean After-Mean Before)] – [In individual Clinician-focused (Mean After-Mean Before)]
<sup>b</sup> Least squares mean
<sup>c</sup> Part A was the on-line module
<sup>d</sup> Part B was the role-play session

### Qualitative results

Fourteen facilitators were identified across seven of the 14 TDF domains, while 15 barriers were mapped onto nine of the TDF domains. The predominant barriers and facilitators were consistently associated with *environmental contexts and resources* domain, with five barriers and six facilitators falling under this category. Furthermore, the second most prevalent facilitator was linked to *skills* domain, while the second most common barrier was attributed to *social influences* domain. Looking more in depth into the themes that emerged, the most common theme among the barriers and facilitators in both arms was having enough time to have serious illness conversations. Having an organized workflow appeared to be an important facilitator in the team-based arm, but not in the individual clinician-focused arm. Similarly, communication issues with patients and their own discomfort with serious illness conversations were more frequent barriers with participants in the team-based arm than in the clinician-based arm. At the same time, the team-based approach seems to have added barriers related to interprofessional coordination and interprofessional communication that were not an issue in the individual clinician-focused arm (Table 6).

**Table 6:**
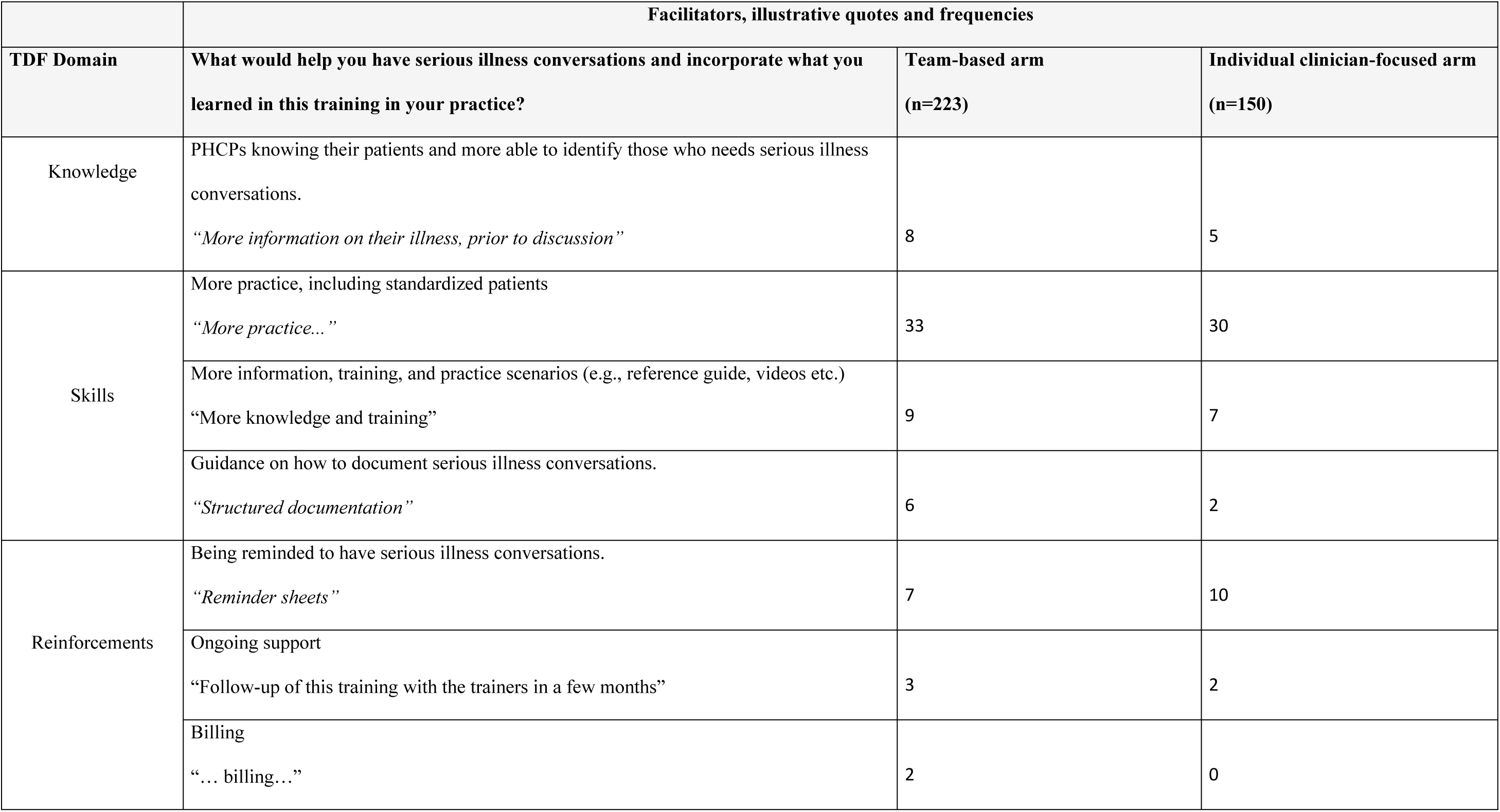
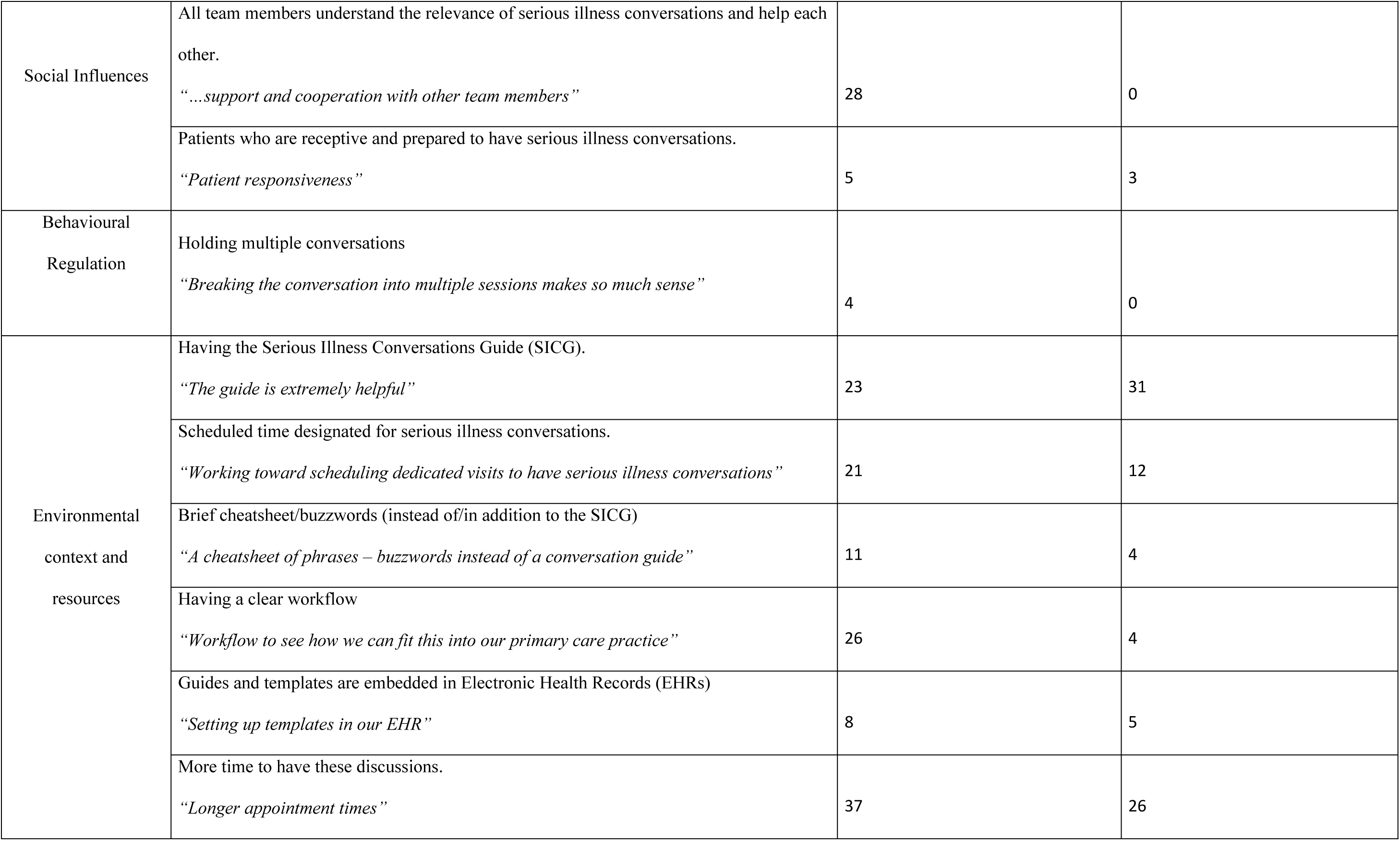
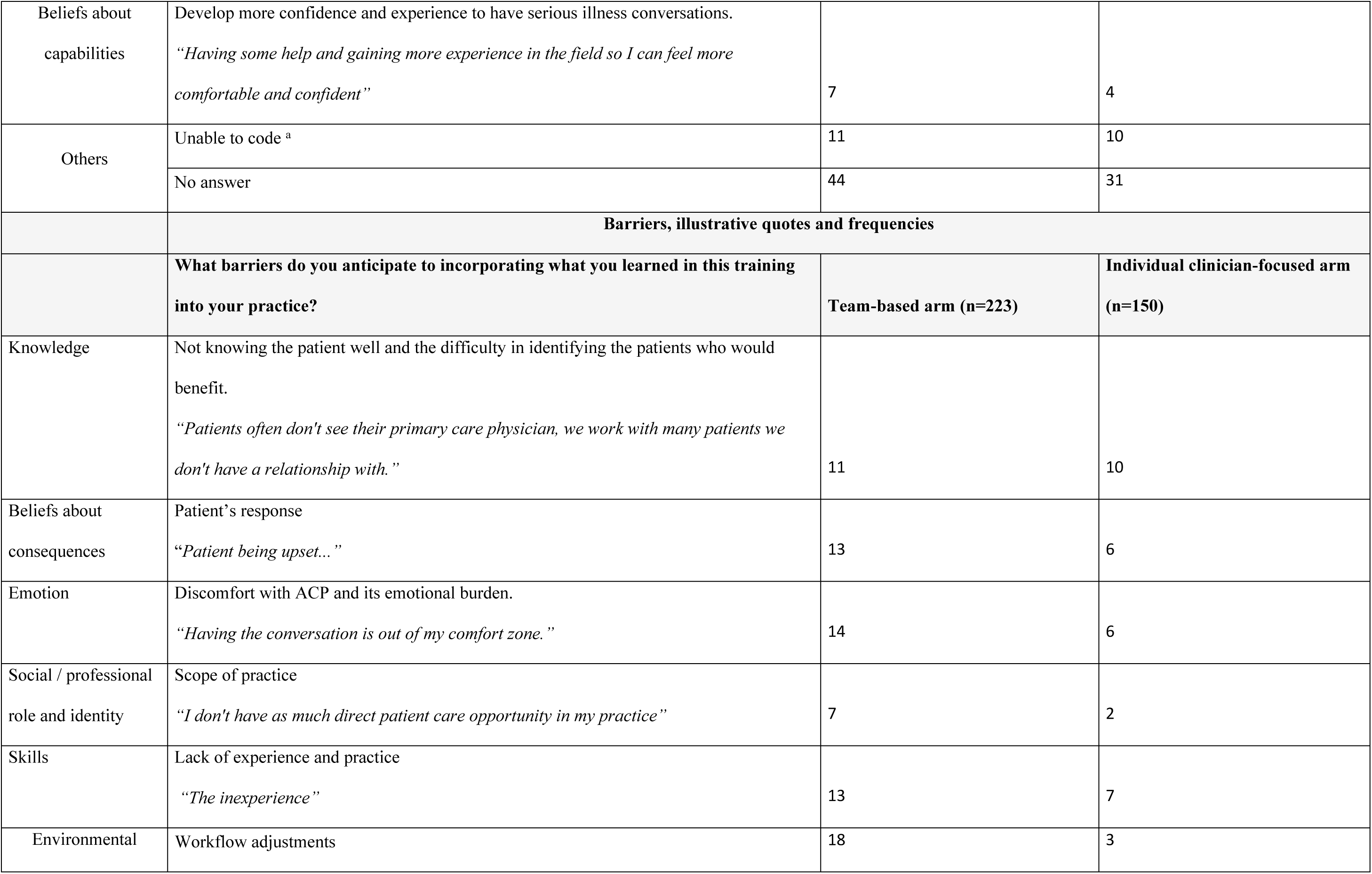
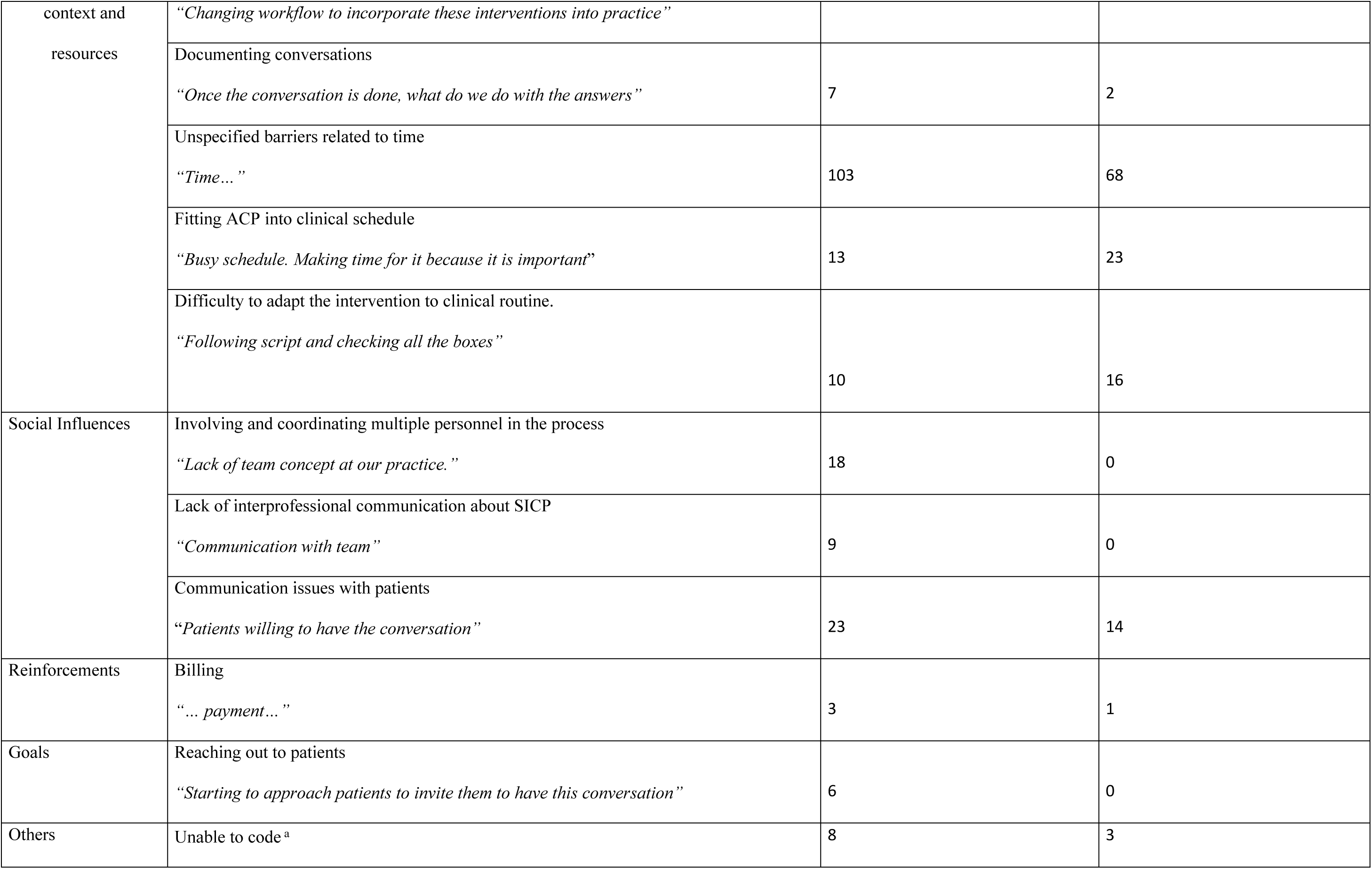
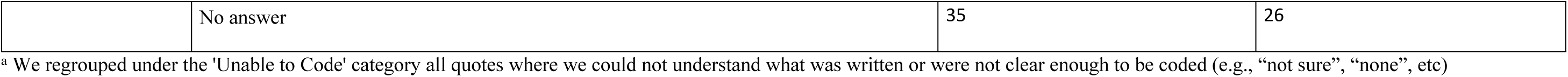
Mapping facilitators and barriers to the Theoretical Domains Framework (TDF) with illustrative quotes and frequencies by arms.

### Triangulation

The psychosocial factors associated with CPD-Reaction matched with the TDF domains of beliefs about capabilities, beliefs about consequences and social influences. Within the TDF, we identified 8 additional psychosocial variables: knowledge, skills, reinforcements, behavioural regulation, environmental context and resources, emotion, social/professional role and identity, and goals. The most frequent barrier and facilitator in both arms (environmental context and resources) was classed into the “opportunity” component of COM-B. Concurrently, comparative analysis of the arms using the COM-B revealed a large portion of the barriers for the team-based arm derived from the domains of “opportunity” (social influences) and “motivation” (emotion and social/professional role and identity). Triangulating the results led to comprehensive recommendations (S2 Table). To enhance the intervention, suggestions were grounded in behaviour change techniques linked to functions such as Education, Modelling, Enablement, Environmental Restructuring, Training, and Persuasion. Notably, Environmental Restructuring (n=7) and Education (n=5) emerged as the most frequently recommended functions (S2 Table)

## DISCUSSION

We conducted a mixed-methods theory-driven process evaluation alongside a cRT that compared the impact on patients of a team-based SICP training approach for PHCPs with the impact of an individual clinician-focused SICP training approach, with post-intervention measures only. Our process evaluation explored the causal mechanisms in operation, specifically relating to the PHCPs taking the training, that may have influenced the success or not of the intervention, more specifically their intention to have serious illness conversations with patients. Contrary to our hypothesis, we found that intention was lower in the team-based (intervention) arm than in the individual clinician-focused arm, but after adjusting for age, profession and education this difference was not statistically significant. We also found that likelihood to engage in ACP before training (assessed retrospectively) was lower in the team-based arm than in the individual clinician-focused arm. Both arms assessed likelihood to engage patients in ACP and to use the SICG as higher after training than before, but the mean difference between the likelihood before and after the training was no higher in the team-based arm than in the individual clinician-focused arm. Environmental context and resources were the most common TDF domains that emerged from the barrier and facilitators questions. Lack of time was a major barrier in both arms, while organized workflow was a key potential facilitator in the team-based arm. Additionally, the team-based arm reported more emotional discomfort and concerns about patients’ reactions to serious illness conversations, as well as challenges coordinating multiple personnel and communicating interprofessionally, that mapped onto the *social influence* TDF domain. The latter correlates with the lower mean on the team-based arm in the social influences CPD-Reaction construct. By triangulating results using the COM-B framework, we found that most barriers and potential facilitators for both arms related to the *opportunity* domain. The COM-B model informed our theory-based recommendations, leading to the identification of areas for improvement in future interprofessional SICP interventions. These recommendations primarily focused on environmental restructuring. These results lead us to the make the following observations:

First, contrary to our main hypothesis, the team-based arm had a lower level of intention to have serious illness conversations with patients than those in the individual clinician-focused arm. However, the difference was no longer statistically significant when adjusted for age, profession and education. To the best of our knowledge, this study is the first study to measure PHCPs’ intention to have serious illness conversations with patients. Therefore, we cannot directly compare our results with those of other studies. Yet, related evidence on ACP programs tends to favor team-based approaches (13, 53). The differences in the professions found in each arm of the parent cRT may explain these findings. Our process evaluation also suggested that the other primary care team members participating in the team-based arm may not, in their practices or healthcare systems, have had the deontological responsibility or the expectations to have this type of discussion. Qualitative data support this assumption since it showed that in the team-based arm more PHCPs perceived serious illness conversations as outside their scope of practice. Practices vary between countries, states, and provinces regarding integrating palliative care practices into primary care, and especially regarding who should initiate serious illness conversations (11, 54, 55). For some of the other primary care team members this may have been the first time they considered engaging in such conversations (56). In a survey of interprofessional healthcare professionals in Colorado, most agreed that it was exclusively the physician’s role to discuss ACP with patients and family (57). Future initiatives to improve the training should focus on integrating a more comprehensive approach, fostering consistent support from managers or team leaders and using the expertise and assistance from more experienced PHCPs as an effective strategy to improve the sense of ownership regarding conversations and consequently bolster the intention of those less experienced to have serious illness conversations with their patients (54, 55).

Second, PHCPs in both arms assessed their likelihood to engage in ACP before training as low, and the team-based arm rated it even lower than the individual clinician-focused arm. This could indicate that at study outset, baseline intention was low in the team-based arm, supporting our assumption that the lack of experience of some of the other primary care team members of the team-based arm may have affected results. However, in both arms the training appeared to increase the likelihood of engaging in ACP and using the SICG, suggesting an overall positive impact of the SICP training sessions using either approach. A 2019 literature review on ACP in multiple settings also showed an overall positive effect of training on healthcare professionals’ comfort with discussing end-of-life decisions (58). Similarly, equipping PHCPs with the tools to have end-of-life discussions, such as the SICG, proved to be positive, as shown by the likelihood to use the SICG. Still, as evidenced by our qualitative data, there is a great interest in other types of tools such as cheatsheets and EHR embedded tools. The findings are further corroborated by research conducted in other settings, where tools supplementing the SICG (e.g., SIC reminders, behavioural nudges) emerged as crucial elements in facilitating successful implementation (59–62). Consequently, future interventions adapting the SICP to a primary care context should prioritize equipping PHCPs with a wider range of tools beyond the SICG. These additional tools should be readily embeddable within their existing clinical practice routines, thereby enhancing their capacity to have effective serious illness conversations.

Third, most frequent barriers and facilitators raised by PHCP’s were related to the environmental context. For example, time emerged as both a barrier and a facilitator in both arms. We expected a team-based approach would reduce the time PHCPs need for discussions by promoting the sharing of tasks. It is possible that team-based SICP still lacks a better method of sharing tasks to be effective. A study that adapted SICP to other contexts indicated that a well-organized division of tasks and workflow is an essential element for implementation of team-based approaches (61, 63). Our study validated this: participants indicated that team-based training needed to better help teams organize their workflow and clarify and recognize the role of each professional. It is also possible that the lack of effect observed on the intention in the team-based approach could also be attributed to the additional time and burden required for coordination, and communication among team members, as seen in our qualitative data. This should be taken into consideration in future studies. Future adaptations of the team-based SICP should prioritize on building more relevant skills and designating tasks across roles. In addition, participants in the team-based arm expressed more discomfort about discussing serious illness conversations and had greater concerns about patients’ readiness for such discussions, compared to participants in the individual clinician-focused arm. This finding supports our assumption that despite the training provided to healthcare professionals, there remains a gap in their understanding and preparedness for engaging in serious illness conversations, particularly among the other primary care team members of the team-based arm. This suggests that to ensure effective serious illness conversations, further efforts are needed to adapt the training to address their discomfort, fears of negative reactions and concerns about patients’ preparedness.

Fourth, the CPD-reaction construct “social influences” exhibited the lowest mean score among the psychosocial factors influencing intention measured. This finding aligns with qualitative results from the team-based arm, where relevant barriers such as interprofessional team coordination and communication difficulties were associated with the corresponding TDF domain. “Social influence” is defined as the perception of approval or disapproval from significant peers or social referents regarding the adoption of a specific behavior (36). Based on this definition, one might expect PHCPs participating in team-based training to report higher levels of social influence compared to those in individual training. However, our study did not find this to be the case. Still, this finding aligns with existing research on the relationship between social influence and behavioural intention. A systematic review of 52 studies revealed that social influence had the lowest average score range among the four psychosocial factors influencing intention (64). Similarly, a qualitative study using an interprofessional approach mapped social influences onto relevant barriers to behaviour adoption(65) Nevertheless, Michie et al. established the social influence domain as a crucial target for health intervention planning (45), further supported by a systematic review analyzing 277 articles about behaviour change techniques (66). Therefore, despite the interprofessional design of our intervention, enhancing social support measures remains essential. This could include providing more information about the level of peer approval, facilitating understanding and trust through social comparison, and incorporating additional social support/social change strategies (18, 26). Thus, refining implementation strategies related to social influence will potentially increase the success of the team-based training.

Lastly, our data triangulation revealed that the majority of barriers and facilitators to behavior adoption, especially on the team-based arm, stem from the “opportunity” domain of the COM-B framework. This finding aligns with the framework’s conceptual model, which suggests that opportunity influences capacity (encompassing intention), which in turn influences behavior (S2 Fig) (18, 49). Therefore, there remain upstream barriers and potential facilitators, particularly in the environmental context and resources domains, that limit behaviour change. Consequently, in line with the COM-B framework, modifications targeting environmental changes are a way to improve the success of the intervention (28). These findings suggest that more than training, structural adjustments within the clinical setting are crucial for facilitating such conversations. Incorporating these changes and mention them in team-based SICP training itself could offer significant value. However, the wider scope of our study, which encompassed primary care settings in two countries, precluded the use of targeted environmental changes into practices. The context of the practices varied greatly, making it impossible, for example, to integrate a “one-size-fits-all” workflow into the training or incorporate an EHR reminder into all practices. This finding highlights a critical obstacle to large scale interventions: balancing the need for standardized content across diverse clinical contexts with the need to ensure adaptability to each practice’s specific needs (67–69). However, this limitation, combined with a process evaluation, enabled the identification of key SICP components critical for maintaining and replicating its effectiveness (5, 6, 59, 60). As Konnyu et al. emphasize, mapping the potential for adaptation and intervention based on behavior change techniques is crucial for identifying potential gaps and optimizing interventions accordingly (70). Our findings, therefore, support the significance of environmental context modifications as effective tools for behavior change in future SICP implementations, be it in research settings or healthcare policies.

### Limitations

A true pre-intervention measure of baseline intention to engage in serious illness conversations with patients would have been preferable, but pragmatic considerations precluded this. It is difficult to show differences in comparative effectiveness studies in real-world clinical practice, particularly when interventions are similar. (71) Second, this study is a process evaluation alongside a cRT in which randomization units were the clinical practices and not the PHCPs. Although characteristics of the clinical practices were balanced between arms, PHCPs’ characteristics were not balanced as involving different professions was part of the design, therefore the comparison between arms for our main outcome, intention, could be limited. However, we controlled for these variables in the multivariable analysis, and we conducted subgroup analyses of similar professions and, our finding that intention increased more in the individual clinician-focused arm was no longer significant. This study analyzes team and clinician outcomes of a cRT for which the sample size and statistical power were calculated for patient outcomes, as these were the primary outcomes. Participation of PHCPs was encouraged but the main focus was on patient recruitment and the number of practices, not the number of PHCPs. Thus, our results should be interpreted with caution, as their lack of significance may be secondary to lack of statistical power, reducing the precision of our results. Future studies evaluating a team-based approach should therefore increase the number of team participants. Finally, the study’s qualitative data collection method, relying solely on post-training open-ended questions, represents a limitation. While these questions provided valuable insights, the absence of semi-structured interviews or focus groups restricted the data’s depth and complexity. This restriction stems from time constraints in real-world implementation projects. The 3-hour training session had already consumed a substantial portion of this allocated time, precluding the inclusion of more elaborate qualitative data collection methods. Despite this limitation, the open-ended question findings correlated with quantitative data, supporting the study’s conclusions.

## CONCLUSION

Our theory-based process evaluation alongside a cRT showed that when an individual clinician-focused SICP training approach was compared with a team-based training approach, the latter had no more impact on increasing PHCPs’ intention to have serious illness conversations with patients. However, both training programs were useful for increasing PHCPs’ likelihood of engaging in ACP discussions and to use the SICG guide. A team-based approach requires further focus on training the other primary care team members to feel empowered and responsible to have serious illness conversations and to address their fears of negative reactions, and their concerns about patients’ preparedness. Our findings suggest that future adaptations of the team-based SICP training approach should prioritize on building more relevant skills and designating tasks across roles. Finally, our data triangulation revealed that the majority of barriers and facilitators to behavior adoption stem from the “opportunity” domain of the COM-B framework. This suggests that more than training, structural adjustments within the clinical setting are crucial for facilitating serious illness conversations. Overall, our study provides valuable insights into the challenges and opportunities for implementing team-based SICP training in primary care. By addressing these findings, future interventions can improve the effectiveness of SICP training and support PHCPs in having more meaningful, high-quality and timely serious illness conversations with their patients.

## Data Availability

All relevant data are within the manuscript and its Supporting Information files.

## ACKNOWLEDGEMENTS

We acknowledge the precious work of Louisa Blair for her editorial help with the manuscript and Stéphane Turcotte for his vital insights in the statistical analysis. We also thank the members of the Meta-LARC ACP cRT team for their involvement in this project.

## COLLABORATORS META-LARC ACP TRIAL TEAM

Angela K. Combe, Oregon Health & Science University

Annette M. Totten, Oregon Health & Science University

Barcey T. Levy, University of Iowa

Cat Halliwell, University of Colorado

David A. Dorr, Oregon Health & Science University

David Nowels, University of Colorado

Deb Constien, Patient-partner

Deborah Dokken, Patient-partner

Donald E. Nease, Jr., University of Colorado

Dr. B. Angeloe Burch Sr., Patient-partner

Elizabeth Fernley, Oregon Health & Science University

France Légaré, Université Laval - VITAM - Centre de recherche en Santé durable

Gail Drey, Patient-partner

Gurnoor Kaur Brar, University of Toronto

Jacqueline D. Alikhaani, Patient-partner

James Pantelas Patient-partner

Jean-Sebastien Paquette, Université Laval - VITAM - Centre de recherche en Santé durable

Jeanette M. Daly, University of Iowa

Jessica E. Ma, Duke University

Jodi Lapidus, Oregon Health & Science University - Portland State University School of Public Health

Judy Katz, Patient-partner

Kate Hanrahan, University of Iowa

Kathy Kastner, Patient-partner

Katrina Ramsey, Oregon Health & Science University

Keith Provin, Patient-partner

Kirsten Wentlandt, University Health Network

Kylie Lanman, Oregon Health & Science University

LeAnn C Michaels, Oregon Health & Science University

Lyle J. Fagnan, Oregon Health & Science University

Mary F. Henningfield, PhD, University of Wisconsin-Madison

Mary M. Minniti, Patient-partner

Matthew Howard, Oregon Health & Science University

Megan Schmidt, University of Iowa

Meredith K. Warman, University of Colorado

Michelle Greiver, University of Toronto

Olga Petrova, Patient-partner

Patrick M. Archambault, Université Laval - VITAM - Centre de recherche en Santé durable and Centre intégré de santé et de services sociaux de Chaudière-Appalaches

Peter Kim, University of Iowa

Rowena J. Dolor, Duke University

Sabrina Guay-Bélanger, VITAM - Centre de recherche en santé durable

Sarah Bumatay, Oregon Health & Science University

Sarina Schrager, University of Wisconsin-Madison

Sean Rice, Oregon Health & Science University-Portland State University School of Public Health

Sharon E. Straus, University of Toronto

Shelbey Hagen

Shigeko (Seiko) Izumi, Oregon Health & Science University

Souleymane Gadio, VITAM - Centre de recherche en santé durable

Suélène Georgina Dofara, VITAM - Centre de recherche en santé durable

Susan Lowe, Oregon Community Health Information Network - Columbia Gorge Health Council

Taryn Bogdewiecz, University of Colorado

## Supporting Information

**S1 Fig. Diagram of the measured behaviours and their interaction**

**S2 Fig. The COM-B model and its correlation with the TDF domains**

**S1 Table. ACP engagement scores before and after training**

**S2 Table. Recommendations to improve the training based on barriers and facilitators, using the COM-B model, the Theoretical Domains Framework and the CPD-Reaction questionnaire.**

**S1 Checklist. CONSORT**

**S2 Checklist. GRAMMS**

## Notes

### Competing Interest Statement

The authors have declared no competing interest.

### Clinical Trial

ClinicalTrials.gov (ID: NCT03577002).

### Clinical Protocols

https://doi.org/10.1089/jpm.2019.0117

https://clinicaltrials.gov/study/NCT03577002?id=NCT03577002

### Funding Statement

Yes

### Author Declarations

The parent META-LARC ACP cRT was approved by the Trial Innovation Network Single IRB at Vanderbilt University Medical Center (IRB#181084) for the U.S. sites by the Research Ethics Board of the Centre Intégré Universitaire de Santé et de services sociaux (CIUSSS) de la Capitale-Nationale in Quebec City, Canada (ethics number #MP-13-2019-1526), for the sites in Quebec and by the Health Sciences Research Ethics Board of the University of Toronto (protocol number 36631), for the sites in Ontario. The process evaluation's data and outcomes were included in the approval. All subjects willingly agreed to take part in the study, and their consent was obtained in accordance with the regulations of the Institutional Review Board or Research Ethics Board in effect.

